# COVID-19 diagnostic testing underestimated cases amongst females in Pakistan

**DOI:** 10.1101/2023.01.30.23285225

**Authors:** Najia Karim Ghanchi, Kiran Iqbal Masood, Asghar Nasir, Nazneen Islam, Zeeshan Ansar, Zahra Hasan

## Abstract

**Objectives:** Understanding the impact of COVID-19 largely depended on information from PCR based diagnostic testing of SARS-CoV-2. It was recognized early in the pandemic that testing rates varied greatly between high and low income countries. Whilst total numbers of tests conducted are noted, little attention has been made to differences that may be due to gender and we examined this in the context of Pakistan.

**Methods:** We conducted a retrospective cross-sectional analysis of respiratory specimens received for SARS-CoV-2 by PCR at Aga Khan University Hospital, Karachi, Pakistan between February 2020 and February 2022. Data was analysed in six monthly intervals; P-I, February to July 2020; P-II, August 2020 to January 2021; P-III, February to July 2021 and August 2021 until February 2022.

**Results:** A total of 470,047 PCR tests were conducted. The proportion of tests conducted for females was, 35% in P-I; 40% in P-II; 44% in P-III and 46% in P-III. 21% of specimens were positive for SARS-CoV-2, only 9% of these specimens were from females. The greatest numbers of tests were conducted in males aged 31 to 45 years followed by those aged 16-30 years. The fewest tests were conducted in children aged under 15 years. The highest percentage of PCR positive tests was found in those ages 60 years and above. Compared for gender SARS-CoV-2 positivity rates were comparable across the study period.

**Conclusions:** COVID-19 data from Pakistan indicates that there are larger numbers of males as compared with females who were affected by this disease. Our results show that this may be due to a gender bias in the demographics of testing. This was especially true in the early pandemic period, leading to under-surveillance and -reporting of COVID-19 cases in females especially, in those of younger and older age groups.

**Strengths and limitations of this study:**

- we stratified PCR testing based on gender and provide information on diagnostic trends
- we found COVID-19 rates were comparable between females and males
- Fewer samples from females were received particularly, in 2020 with an increase by 2022
- it proposes the importance of examine the total samples tested in any case before drawing conclusions based on results of laboratory reports.

**Limitations:** This study was from one private healthcare system and cannot completely reflect nationwide trends to limited access to testing and also stigma related to the COVID-19.

## Introduction

The global outbreak of a novel coronavirus causing severe acute respiratory distress syndrome 2019 (COVID-19) was declared a Public Health Emergency of International Concern on 30 January 2020 (1) and a pandemic on March 11, 2020, by the World Health Organization (WHO). Countrywide data for Pakistan informs us that to date 1.58 million COVID-19 cases have been reported with a mortality of 30,639 (up until 13 January 2023). Globally, it is data from SARS-CoV-2 PCR tests of respiratory samples that have informed us of COVID-19 transmission patterns, leading to an understanding of disease progression and severity, and providing insights into public health management of the disease.

Public health laboratories across Pakistan worked together with private laboratories and hospitals to provide diagnostics for COVID-19. The Aga Khan University Hospital (AKUH), Karachi, Pakistan was at the front-line of COVID-19 testing since February 2020. Diagnostics for COVID-19 were challenging to introduce and scale up during the first year of the pandemic. This was particularly difficult during the first six months of the pandemic when lockdowns were in place making it difficult to import testing reagents into Pakistan. 135,766 COVID-19 cases had been reported from Sindh province by September 2, 2020; of these, 30% were females and 70% were males (3). There is limited information regarding to the pattern of COVID-19 testing across the pandemic period in Pakistan. Here we have examined gender and age-wise trends of specimens received for SARS-CoV-2 at AKUH, comparing trends in six monthly intervals during the period February 2020 until February 2022.

## Methods

This was a retrospective cross-sectional analysis of clinical laboratory records at the Aga Khan University Hospital, Pakistan,

Nasal/ Nasopharyngeal swab specimens were tested for SARS-CoV-2 using the Cobas^®^ SARS-CoV-2 RT-PCR assay (Roche Diagnostics, USA). Here we included data for specimens submitted in the province of Sindh. PCR testing data was analysed in six monthly intervals; P-I, February to July 2020; P-II, August 2020 to January 2021; P-III, February to July 2021 and August 2021 until February 2022.

Demographic data including age and gender, and clinical and laboratory information was documented. Data was analyzed using SPSS (Social Science Statistical Software Package).

Patients and Public Involvement did not occur for this manuscript.

## Results

Our laboratory conducted 470,047 PCR tests during the study period February 2020 until February 2022. Five COVID-19 waves were observed during this time as depicted in Figure 1A. Overall, 21% (96,977) of all tests were positive for SARS-CoV-2; comprising 9% of specimens from females and 12% from males. We examined pattern of COVID-19 diagnostic testing in six monthly periods; February – July 2020, P-I; August 2020 – January 2021, P-II; February – July 2021, P-III and August 2021 and February 2022, P-IV. Of the nasal swab specimens received for testing, those that belonged to females were fewer than from males in each of the periods; P-1 to P-IV (Figure 1B) with tests for females representing, 35%, 40%, 44% and 46% respectively of total samples teste during the intervals. The SARS-CoV-2 positivity during P-I, P-II, P-III and P-IV was 33%, 18%, 18%, and 21% respectively. For females, it was found to be 32%, 19%, 18% and 20%, Figure 1C. Notably, the PCR test positivity was found to be comparable between females and males during each time interval studied.

**Figure 1.**
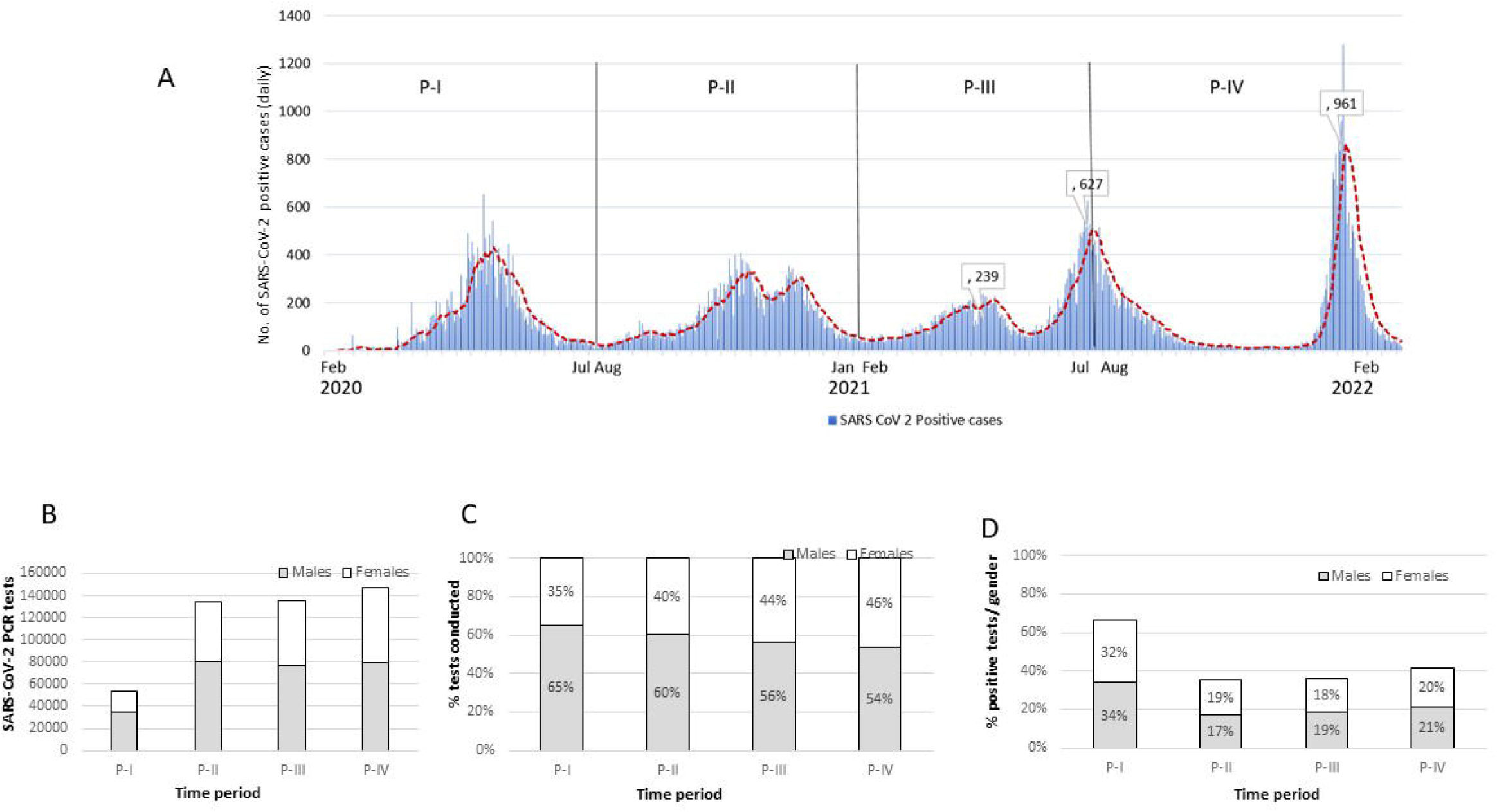
Trend of COVID-19 positive cases reported from Aga Khan University Hospital. The figure depicts the number of daily cases reported as SARS-CoV-2 positive by AKUH, Clinical Laboratories for the period February 2020 until February 2022 (A). This period was divided into six monthly intervals February 2020 – July 2020 (P-I), August 2020 – January 2021 (P-II), February 2021 – July 2021 (P-III) and August 2021 – February 2022. Graphs show the number of respiratory specimens received for SARS-CoV-2 PCR testing for males and females (B), and the proportion of males and females who were PCR positive when determined separately for each gender (C).

We further investigated if there was an age-wise trend in the PCR testing conducted. The mean age of COVID-19 cases was 39 ± 17 years. When we looked at both age and gender together, it was males aged 31 to 45 years followed by those aged 16-30 years for whom the largest number of tests were conducted, Supplementary Table 1. The greatest number of SARS-CoV-2 positive individuals were those aged 31-45 years, 21% of total tested in this group, the largest number belonging to males, Supplementary Table 1. The fewest PCR tests were conducted in children aged under 15 years and under (16%), Supplementary Table 1. The highest percentage of PCR positive tests was found in those ages 60 years and above.

To understand the testing patterns over the 2 year study period, we compared the proportion of age-wise testing in females and males between February 2020 and February 2022. The higher proportion of males tested during P-I was evident across all age groups (p< 0.0001), Figure 2A. Fewer PCR tests were conducted for females aged 31-45 years and those aged 46 – 59 years. Significantly fewer positive PCR tests were reported in females of all age groups during P-I, Figure 2B. During P-II and P-III, the number of tests conducted for each group were reduced for females aged 31-45 years. Similarly, fewer COVID-19 cases were identified in females of this group during P-II and P-III (p< 0.0001). During P-IV, there was no difference in the numbers of respiratory specimens received from females and males. Similarly, the percentage of COVID-19 positive individuals in each group was found to be similar during this period.

**Figure 2.**
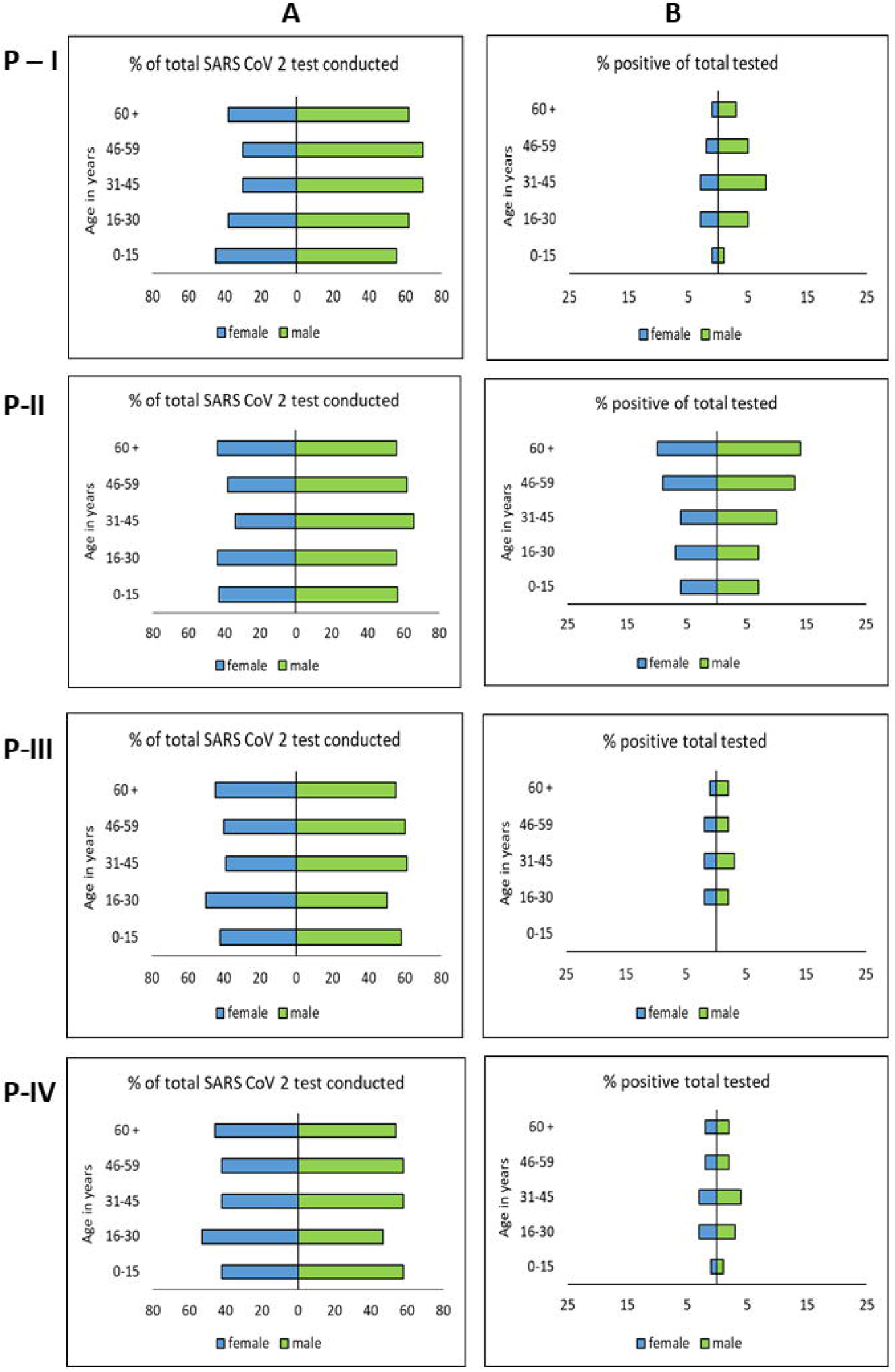
Gender and age-disaggregated data for SARS-CoV-2 PCR tests conducted February 2020 – February 2022. Panels show data in six monthly intervals; P-I, February – July 2020; P-II, August 2020 – January 2021; P-III, February – July 2021 and P-IV, August 2021 and February 2022. The left panels (A) show the percentage of females and males tested within each age group and right panels (B) show the percentage of individuals who tested positive for SARS-CoV-2 within each gender group.

## Discussion

We describe the trends of SARS-CoV-2 PCR testing in Pakistan. Our data highlights limited COVID-19 diagnostic testing for SARS-CoV-2 in females especially, in the first year of the pandemic. Provincial and National data on COVID-19 during the early pandemic period of 2020 reported 70% of cases in males and 30% in females (2, 3). This did not fit with global data as, in 2020, COVID-19 and Gender age dis-aggregated data showed comparable incidence between males and females except in some countries such as, Pakistan, India, Nepal, Bangladesh and Afghanistan, which reported higher confirmed COVID-19 cases in males than females (2, 4).

The number and proportion of women tested for SARS-CoV-2 infection was in the first six months of the pandemic between February and July 2020. This was the time when there was both limited availability to COVID-19 diagnostics but also stigma related to the disease in Pakistan (5). This stigma may have been magnified in women, especially as reports show there is stigma for women associated with the diagnosis of other infections such as, tuberculosis and breast cancer (6, 7). Later, COVID-19 testing amongst females increased between August 2021 and February 2022, females were amongst 46% of all cases tested. This may be due to the increased knowledge available about COVID-19 at this later time, with diagnostics more commonplace and accessible. Further, the lower number of females represented amongst tests conducted could be due to limited access due to social and financial constraints. In 2020, a study on the possible gender impact and implications of COVID-19 in Pakistan highlighted that women could be disadvantaged due to their inequalities in the context of literacy, financial empowerment and access to health care (8).

Overall, we found the greatest number of SARS-CoV-2 PCR tests were conducted in those aged between 16 and 45 years of age. This fits with the demographics of the population whereby, 69% of the population is under 30 years whilst 6% are greater than 60 years of age (9). National data on COVID-19 reports indicates that 76% of confirmed cases were below 50 years old (3).

When we looked at both age and gender together, it was males aged 31 to 45 years followed by those 21 to 30 years which formed our largest cohort of COVID-19 cases. Males of this age group are more mobile, comprise the larger part of the workforce and are likely to access testing. Whilst, the largest proportion of positive cases/age group was in those aged 60 years and above. This has been shown to be the highest-risk age group for COVID-19 as males are shown to be more susceptible to severe disease than females (10, 11).

Females aged below 15 years and those 60 years and above were the group least tested for COVID-19. It may be that they had decreased mobility and access to testing. Health care initiatives were shown to be impacted for older women during the COVID-19 pandemic (12). Globally, the COVID-19 pandemic has exposed inequalities in social and economic impact on women (13). In Pakistan, health-seeking behavior is affected by socio-economic factors, literacy and gender bias (14, 15). For COVID-19, it is likely that fewer females either volunteered to get tested or had the opportunity to seek testing. This indicates a gender-bias in testing which may have led to under-surveillance of females with COVID-19 in Pakistan.

## Data Availability

All data produced in the present work are contained in the manuscript

## Contributorship statement

ZH designed the study. AKG, KIM, AN, NA and ZA were involved in validation and methodology. NKG and ZH wrote the first draft. All authors approved the final draft. We thank Kausar Hamid and Kahkashan Imam for technical support provided in this work.

## Acknowledgements

We thank the Department of Pathology and Laboratory Medicine, Aga Khan University, Pakistan and the Clinical Laboratories and their collaboration with Department of Health, Sindh to establish COVID-19 testing services during the pandemic.

## Data sharing statement

Data pertaining to the study will be available upon reasonable request after the publication of this manuscript.

## Ethics approval statement

This study was approved by the Ethical Review Committee, Aga Khan University, Pakistan. It was an exemption study that did not involve any personal data from individuals.

