## Supplemental Table for "COVID-19 diagnostic testing underestimated cases amongst females in Pakistan"

**Supplementary Table 1. Gender distribution of respiratory specimens received for SARS-CoV-2 PCR testing**

|  | PCR tests | Total tested | | % tested within group | | SARS-CoV-2 positive |  | SARS-CoV-2 positive (n) | | % SARS-CoV-2 positive | | % SARS-CoV-2 positive of total group | |
| --- | --- | --- | --- | --- | --- | --- | --- | --- | --- | --- | --- | --- | --- |
| Age groups | (n) | females | males | females | males | number | %/total | females | males | females | males | females | males |
| Total | 470047 | 198771 | 271276 |  |  | 96977 | 21 | 40243 | 56734 | 20 | 21 | 9 | 12 |
| 0-15 | 34062 | 14592 | 19470 | 43 | 57 | 5369 | 16 | 2478 | 2891 | 17 | 15 | 1 | 1 |
| 16-30 | 145196 | 69619 | 75577 | 48 | 52 | 24020 | 17 | 11312 | 12706 | 16 | 17 | 2 | 3 |
| 31-45 | 145037 | 54525 | **90512** | 38 | **62** | **30836** | **21** | 11671 | **19165** | 21 | 21 | 2 | 4 |
| 46-59 | 82582 | 32134 | 50448 | 39 | **61** | 20246 | 25 | 7856 | 12390 | 24 | 25 | 2 | 3 |
| 60above | 63170 | 27901 | 35269 | 44 | 56 | 16506 | 26 | 6926 | 9580 | 25 | 27 | 1 | 2 |
